# Health-Related Effects of Real-Time Circuit Tele-Training and Gym Resistance-Aerobic Training in Ambulatory Adults with Cerebral Palsy

**DOI:** 10.1101/2024.03.20.24304574

**Authors:** Ronit Aviram, Yisrael Parmet, Simona Bar-Haim

## Abstract

**Objective:** To compare the impacts of gym training and circuit Zoom-tele-training on health parameters in ambulatory adults with spastic cerebral palsy.

**Methods:** Participants were divided into three groups: The Gym-group that underwent resistance-and-aerobic training program (n=12), the Zoom-group that underwent a circuit Zoom-tele-training program (n=14), and the Control-group that was on a waitlist and underwent no training (n=14). The two training gropes exercised bi-weekly for 12-week. Measurements included blood pressure, waist circumference, BMI, 15-repetition maximum-strength tests, and a LALA aerobic shuttle test analyzed using a linear mixed model.

**Results:** Weight circumference decreased in both the Gym and Zoom groups (P=.0202 and P=.0014 respectively); in addition, in both these groups systolic (P=.018, P=.0001 respectively) and diastolic (P=.086 -marginal significance, P<.0001 respectively) blood pressure decreased, with a more pronounced reduction in the diastolic blood pressure for the Zoom-group (P=.043). Maximum aerobic speed increased (P<.0001) in the Zoom-group, with the Gym-group achieving the same speed with a lower peak heart rate (P=.0144). Strength significantly improved in the Zoom group for row (P=.05) and knee-extension (P<.0001) exercises. The Gym group improved in all strength measures (Row P<.0001, Chest-press P<.0001, and Knee-extension P<.0001). The Gym-group’s gains were greater than the Zoom-group’s in the row (P<.0001) and knee-extension (P=.005) exercises. The Control-group experienced a rise in BMI (P=.0256), waist circumference (P=.056 marginal significance), and systolic blood pressure (P=.055 marginal significance).

**Conclusion:** Both exercise programs effectively reduced health-risk factors. The Zoom-group excelled in improving aerobic capacity and diastolic blood pressure, while the Gym-group demonstrated superior strength gains. Not exercising was detrimental to body mass, waist circumference, and blood pressure.

**Impact:** Exercise programs enhance long life heath and prevent health deterioration in adults with cerebral palsy. Results endorse using waist circumference and blood pressure measures as valuable clinical outcomes for adults with cerebral palsy.

## INTRODUCTION

Cerebral palsy (CP) is a group of non-progressive childhood onset motor impairments significantly impacting movement and posture development, thus leading to activity limitations.^1^ As individuals with CP age, they confront an increased risk of developing cardiometabolic health conditions, including type 2 diabetes, hypercholesterolemia, hypertension, excess adipose tissue, and obesity. They also face elevated risks of stroke, cardiovascular morbidity, and mortality, coupled with a high prevalence of chronic multimorbidity.^2–7^

The reduced activity levels in individuals with CP may hinder the generation of muscle force and aerobic fitness beyond the activity threshold, resulting in a reciprocal adverse effect on functional fitness and increased inactivity.^3,8,9^ While low physical activity (PA) levels and poor fitness are recognized risk factors for chronic diseases, incorporating exercise into daily routines holds the potential to enhance fitness and health, breaking the cycle of deconditioning.^8–12^, Existing evidence suggests aerobic training can improve aerobic fitness,^12–14^ and progressive resistance training (PRT) effectively increases muscle generation force (MGF) in individuals with CP.^12,15–17^

Blood pressure (BP), body mass index (BMI), and waist circumference (WC) serve as accessible indicators for monitoring health.^10,11^ The positive effects of exercise on reducing BP and improving body composition are well-documented in various adult populations.^10,11^ Furthermore, PA has been associated with BP and WS in adults with CP.^4^ However, studies investigating the influence of exercise on health conditions in adults with CP are scarce, with limited exploration into the impacts on body anthropometrics or composition.^18^ and a lack of studies on the influence on BP.

Although exercise is recommended for adults with CP, challenges arise; young adults in our focus group cited inaccessibility of community facilities and a lack of necessary accommodations and professional consultation.^19^ Moreover, diminishing physical therapy treatments over the years add to the barriers.^20^ To address these challenges, we designed and implemented a combined aerobic-and-resistance training program in a community gym (G-group) and a real-time circuit tele-training program delivered on Zoom (Z-group). Both programs offered exercising opportunities, the latter complying with Covid-19 restrictions on social gatherings.

While a real-time tele-training program has not been previously conducted with participants having CP, notably, telerehabilitation studies demonstrate effectiveness, safety, and feasibility for individuals with different physical impairments,^21,22^ and real-time Zoom exercise is deemed safe and feasible for elderly adults.^23^ Additionally, circuit training studies, particularly one on young children^24^ and another on home-based circuit training for children and adolescents with CP,^25^ showed improvement in strength and muscle power but mixed results for functional mobility and strength. Our prior research indicated the effectiveness of circuit PRT in improving functional mobility in adolescents and young adults with CP (aged 14–21 years),^26^ yet none measured the influence of circuit training on aerobic parameters, BP, and body anthropometrics.

Our study aimed to examine the impact of combined resistance-and-aerobic training in a gym and a real-time circuit tele-training program delivered on Zoom on health-related factors. We compared these two training methods with a waitlist control group, focusing on BP, body anthropometrics (BMI and WC), and health-related fitness (aerobic and strength). Given current knowledge, we hypothesized that both exercise programs would positively impact fitness, BP, and body composition in the two training groups, expecting no change in the control group. Furthermore, we hypothesized that gains would be greater in the group training in the gym.

## METHOD

### Study Design

This study follows a two-arm controlled (non-randomized) trial with parallel groups. Testers were blind to participants’ group allocation. Approval was obtained from the Ethics Committee of Shamir Medical Center (Assaf Harofeh), affiliated with Tel Aviv University, Israel. The trial is registered in the Israeli database MOH_2018-02-14_002188, and participants were provided informed written consent.

### Inclusion Criteria

Eligible participants were male and female adults with spastic CP, classified under Gross Motor Function Classification System (GMFCS) levels I-III^27^, meeting the following specific criteria: (1) ability to household ambulate; (2) aged between 20 and 50 ±1 years; (3) capability to comprehend basic written instructions; (4) ability to communicate face-to-face and through tele-interaction; and (5) basic self-management skills.

### Exclusion Criteria

Exclusions applied to individuals with degenerative neurological, neuropathic, or muscular diseases, post-polio syndrome, rheumatic diseases, chronic alcoholism, drug abuse, use of β-blocker drugs, unstable seizures, expected changes in medication during the study, orthopedic or neurological surgery within 12 months, or Botulinum Toxin injections within three months prior to the intervention.

### Participant Recruitment and Allocation

Recruitment occurred through rehabilitation centers and social networks. Twelve participants were recruited for exercise at a gym (G-group) in the South of Israel. The Z-group and C-group participants (totaling 29 responders) were recruited nationwide, with the last 14 respondents serving as a wait-list control group (Figure 1).

**Figure 1.**
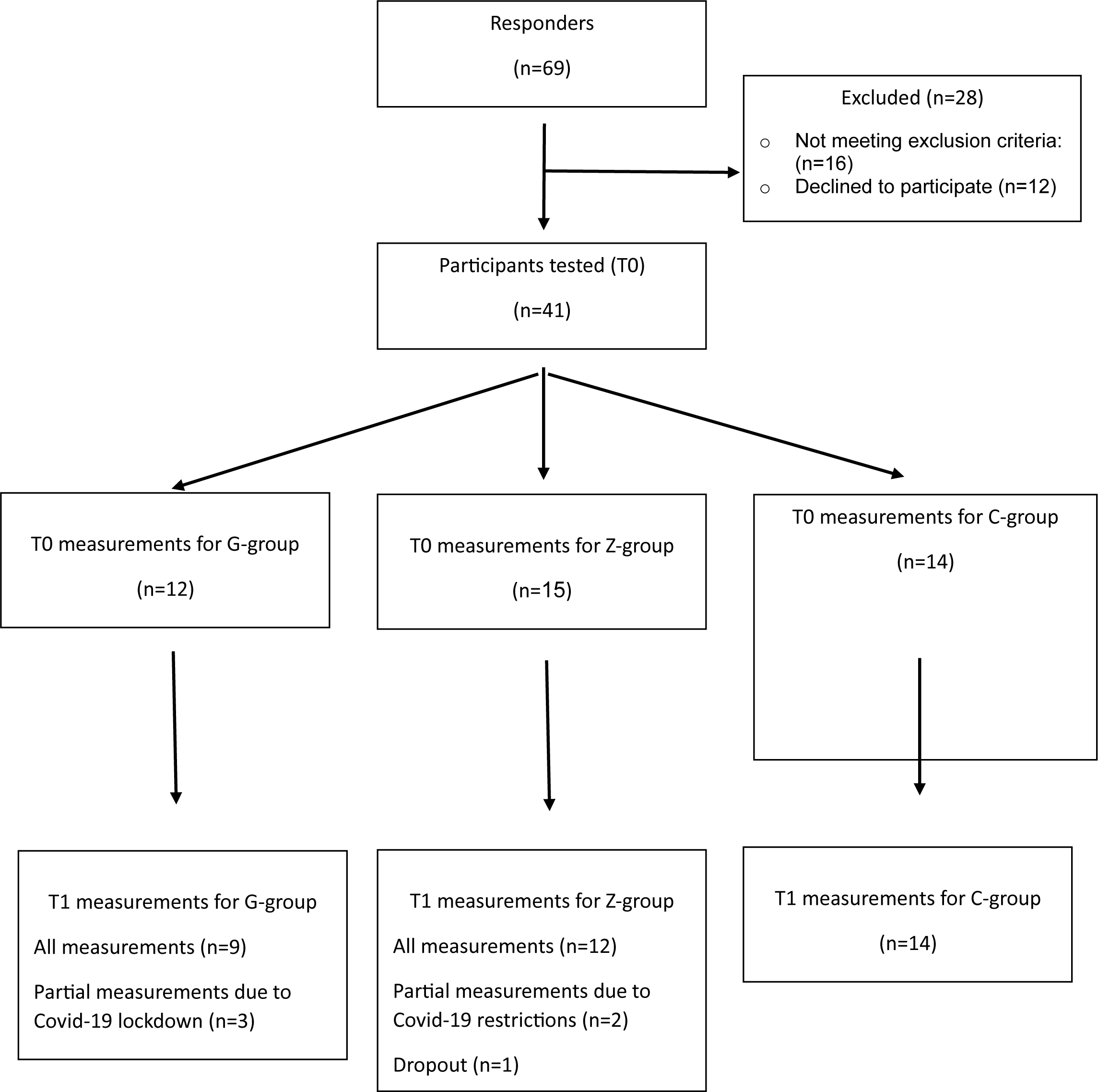
An overview of the patient flowchart

### Interventions

The intervention programs are detailed in the supplementary materials. Briefly, the G-group trained at a community gym, while the Z-group participated in real-time Zoom circuit tele-training at home. Both programs, guided by coaches and physical therapists under the supervision of the first researcher (a physical therapist as well as a PA and sports specialist), entailed 32 one-hour sessions held twice a week; the sessions began with a warmup and ended with a cooldown and stretching. Both training programs were individually tailored, applying the principles of over-load and progression. The aerobic progression plan for the G-group is detailed in Supplementary Tables 1a and 1b; the progression and overload principles are presented in Tables 2a and 2b; and the Z-group’s progression and overload principles are detailed in the text of the supplementary materials. Attention was given to independent and safe training, using an adequate exercise form within personal limits. The G-group underwent aerobic training on a treadmill or combined with another aerobic device. The resistance training included exercises of major muscle groups on the exercise machines (Figure 2). The Z-group training was structured as circuit resistance training comprised of exercises targeting different muscle groups, with quick transitions from one exercise to the next. An illustration of the Z-group training is presented in Figure 3.

**Figure 2.**
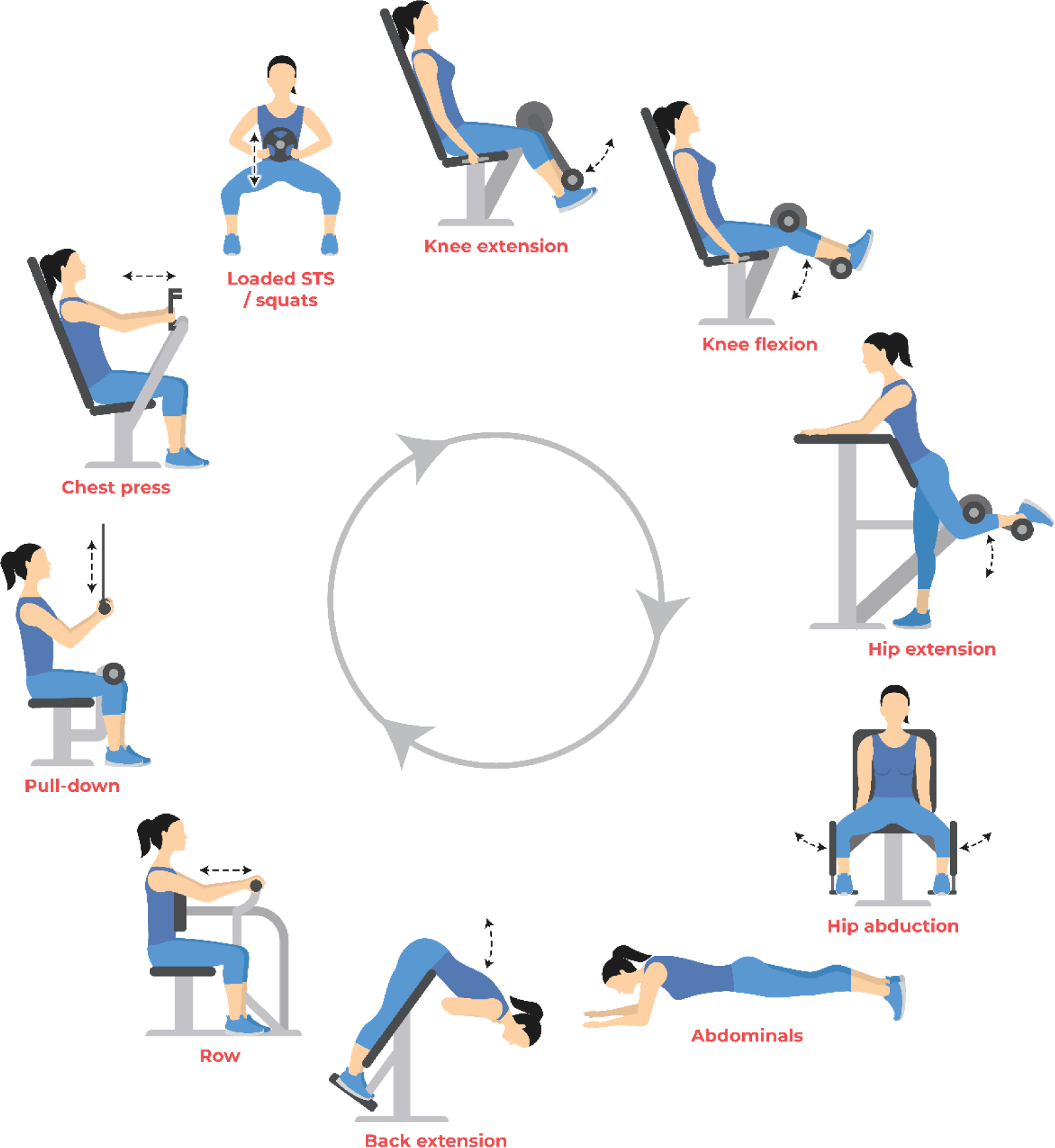
An example of a resistance training exercise in the G-group training, which included the following exercises: knee extension and knee flexions, standing single-leg hip extension, hip abduction, rows, arm pulldowns, chess-press machine exercise, loaded sit-to-stand or squats, back extensions, and abdominal exercise. Each muscle group exercise was offered with different difficulty levels and offers for additional adjustments and adaptations.

**Figure 3.**
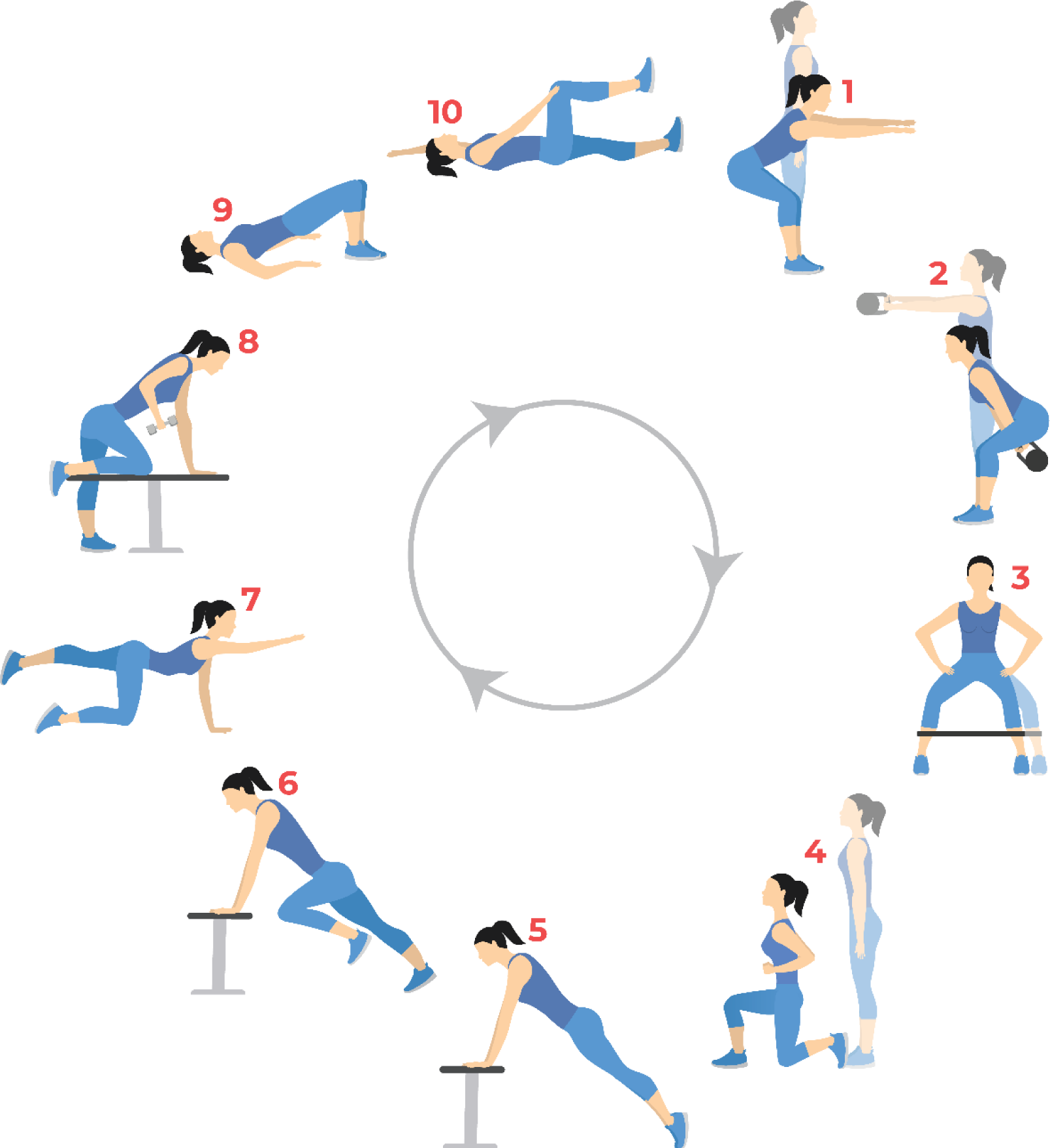
Illustration of the Z-group training: swing (as in the Dumbbell or Kettlebell swing), squats, side Steps to the right and to the left, lunges to the right and to the left, mountain climbers, plank / push-ups, bird dog—two for each hand-leg pair or separate for hands and legs, single arm, chair row, pelvic lift (a.k.a. bridge), and abdominal curl-ups. Each exercise was offered with 3 different difficulty levels and offers for additional adjustments and adaptations.

### Testing and Outcome Measures

Participants in the intervention groups were tested at baseline before training (T0) and three months later at post-test (T1). The C-group was tested at similar intervals without intervention. Testing occurred at the Laboratory of Rehabilitation and Gait Motor Control at Ben-Gurion University of the Negev (G-group) and at Lazus, a community-center gym, in Jerusalem (Z-group and C-group).

The testing order and tester for each participant were maintained throughout the tests. An experienced coach performed the resistance testing, and a physical therapist performed all other tests with the help of an assistant. During the tests, participants employed walking aids and orthoses in accordance with their daily use.

### Outcome measures

#### Anthropometric measures

*Body Mass Index (BMI)* was calculated as the body weight in kilograms divided by height in square centimeters (kg·m−2). Overweight and obese were defined as BMI

≥25kg/m2 and ≥30kg/m2, respectively.^28^

*Waist circumference (WS)* was measured while the participant stood after several consecutive calm natural breaths. A flexible tape measure was wrapped around the participant where the waist is narrowest, the midpoint between the top of the iliac-crest and the lower margin of the last palpable rib. Central obesity was defined as WC ≥80cm for women and ≥94cm for men.^28^

#### Blood pressure

*Systolic blood pressure (SBP) and diastolic blood pressure (DBP)* were measured using a digitally calibrated BP monitor (Omron M6 Comfort HEM-7000-E; Omron Healthcare Co., Ltd., Kyoto, Japan). This monitor demonstrated high validity in adults.^29^ Participants rested, seated with their backs supported for at least 15 minutes before two measurements were taken at 1-to 2-minute intervals, and the lowest values were recorded. Prehypertension is defined as 120–139 mmHg for SBP and 80–89 for DBP, and hypertension as SBP ≥140 mmHg or DBP ≥90 mmHg. In normotensive adults, clinically significant increases in SBP and DBP are 2.4 mmHg and 1.6 mmHg, respectively, in prehypertensive adults, 3.1 mmHg and 1.7 mmHg, and in hypertensive adults 6.9 mmHg and 4.9 mmHg.^11^

#### Fitness

*Perceived fitness level*: Participants were instructed to rate their fitness level as follows: “Rate on a scale of 1-10 how fit you think you are, with 1 representing the least fit and 10 the most fit. Circle the number that best applies: 1,2…. 10.”

*Muscle Strength-15-repetition-maximum (RM)*: 15-RM is the weight an individual can lift for 15 repetitions but no more without compromising the lift. Multiple-repetition testing was suggested as a safe, reliable method.^30^ We used the 15-RM test because of its relevance to our programs. This test was administered by conducting a maximum of two trials for each participant, with at least 5minutes of rest in between.

*Aerobic fitness: The 10-Meter LALA test* was developed by Langerak and Lamberts (Stellenbosch University, South Africa) based on the 10-meter shuttle-walk test.^31^ The 10-meter LALA test is an incremented protocol test paced by a light pacing system (VT-Med, Indico Technologies SRLS, Torino, Italy). The VT-Med system includes an LED strip and control unit that sets the program online (via phone or computer). The test starts at a 2 km/h speed and increases every 20 meters by 0.2 km/h with a second rest, an acceleration of 8%, and a deceleration of 10% every 10 meters to accommodate the participant’s turning around. Failure to maintain the speed of the light at two consecutive turning points determines the end of the test. This uniform protocol enables comparison between participants. HR was monitored continuously using a Tom-Tom HR monitor watch, and peak HR was recorded. This device was found to be reliable and valid.^32^

## SAMPLE SIZE

Preliminary results^26^ indicated that the responsiveness to training in young adults with CP, Cohen’s d^33^ was 0.73. Based on these preliminary results, a calculation of sample size suggested a minimum of 14 participants per group to achieve 80% power with a significance level of 5%. We assumed a potential 20% dropout rate, as observed in our previous study,^26^ and planned the number of participants (51 across all three groups) accordingly.

## STATISTICAL ANALYSIS

The participants’ descriptive characteristics are presented in Table 1. A linear mixed-effects model for repeated measurements was used to test the differences between the two testing periods, baseline (T0) and post-testing (T1), indicating the effects of the interventions. In all dependent variables, the initial fixed factors were the participant group (C-group, G-group, or Z-group), testing time (T0 or T1), and their interactions; the participant factor was taken as a random effect. In all the models, we implemented a process of backward elimination of variables as follows: first, the necessity of the interaction was checked; if it was found to be statistically significant, the process was stopped, and if not, we examined the importance of the two main factors. At the end of model building, the estimated means were calculated for each group and period along with their standard errors, and post-hoc analyses were performed on the factors that were found to be statically significant and remained in the final model. The perceived fitness level scores were measured using a 1-to-10 Likert scale. Therefore, the Wilcoxon rank-sum test with continuity correction was used. All statistical analyses were performed using R-language (R version 4.2.2) with significance assumed at p ≤ 0.05. We used the “lme4” library to estimate the linear mixed-effects models (LMM) and the “emmeans” and “lmerTest” libraries to perform the post-hoc analyses.

**Table 1:** Participants’ characteristics in the three test groups.

| Table 1: Participants' characteristics in the three test groups |  |  |  |  |
| --- | --- | --- | --- | --- |
|  |  | Gym Group<br>(n=12) | Zoom Group<br>(n=14) | Control Group<br>(n=14) |
| Gender<br>(n) | Females | 5 | 10 | 11 |
|  | Males | 7 | 4 | 3 |
| Age: year and month, mean [SD]<br>age range |  | 28y 6.4m [8y]<br>21y 2m-42y-9m | 34y [8y 9.72m]<br>19y 10m-47y 6m | 32y .84m [8y 2.5m]<br>20y 9m-47y 5m |
| GMFCS level (n) | I | 3 | 5 | 2 |
|  | II | 5 | 6 | 9 |
|  | III | 4 | 3 | 3 |
| Type of CP (n) | bilateral limb involvement | 9 | 10 | 12 |
|  | Unilateral limb involvement | 3 | 4 | 2 |

Values are reported as mean [standard deviation] or range. Number of participants (n), Gross Motor Function Classification System<sup>27</sup> (GMFC).

## RESULTS

A summary of the results of the final models obtained for the different dependent variables, the mean estimates and confidence intervals (at a confidence level of 95%), and the P-values are presented in Table 2.

**Table 2:** Heath-related parameters for participants in the Gym, Zoom, and Control groups.

|  |  |  |  |  |  |  |  |  |  |  |  |
| --- | --- | --- | --- | --- | --- | --- | --- | --- | --- | --- | --- |
|  |  | Gym Group |  |  | Zoom Group |  |  | Control Group |  |  |  |
| Est. Mean<br>(95% CI) |  | T0 | T1 | n<br>p-value | T0 | T1 | n<br>p-value | T0 | T1 | n<br>p-value | Time<br>by<br>group-<br>p<br>value |
| Body<br>composition | BMI<br>(kg/m<sup>2</sup>) | 23.40<br>(20.6, 26.1) | 23.00<br>(20.3, 25.8) | 12<br>.65 | 24.90<br>(22.4, 27.5) | 25.00<br>(22.4, 27.5) | 12<br>1.000 | 24.6<br>(22.1, 27.2) | 25.4<br>(22.8, 27.9) | 14<br>.03 | <.01 |
|  | WC<br>(Cm) | 82.5<br>(75.9, 89.4) | 80.50<br>(73.5, 87.4) | 12<br>.02 | 87.20<br>(80.8, 93.6) | 84.60<br>(78.1, 91.0) | 12<br><0.01 | 86.3<br>(79.9, 92.7) | 88.0<br>(81.6, 94.4) | 14<br>.056 | <.0001 |
| Blood<br>pressure | SBP (mmHg) | 121.00<br>(111, 120) | 113.00<br>(104, 122) | 12<br>.02 | 132.00 (124,<br>140) | 120.00<br>(112, 120) | 12<br><.0001 | 129<br>(122, 137) | 135<br>(128, 142) | 14<br>.055 | <.0001 |
|  | DBP (mmHg) | 72.2<br>(65.6, 78.7) | 68.00<br>(31.4, 74.6) | 12<br>.089 | 84.50<br>(78.4, 90.6) | 73.80<br>(37.5, 80.0) | 12<br><0.01 | 81.6<br>(78.6, 87.7) | 85.1<br>(79.1, 91.2) | 14<br>.51 | <.0001 |
| Heath-related fitness | LaLa peak<br>speed (km/hr) | 5.31<br>(4.30, 6.32) | 5.57<br>(4.49, 6.66) | 9<br>.99 | 5.36<br>(4.50, 6.22) | 7.60<br>(6.71, 8.49) | 12<br><.0001 | 5.05<br>(4.03, 6.07) | 5.38<br>(4.37, 6.40) | 14<br>.92 | <.0001 |
|  | LaLa peak HR<br>(bpm) | 152<br>(139, 166) | 141<br>(127, 156) | 9<br><.0001 | 171<br>(158, 184) | 165<br>(152, 178) | 12<br>0.1413 | 165<br>(151, 178) | 167<br>(154, 180) | 0.556 | 0.0714 |
|  | 15RM Row<br>(Kg) | 16.50<br>(9.95, 23.0) | 40.20<br>(33.6, 46.7) | 12<br><.0001 | 23.6<br>(17.6, 29.7) | 30.00<br>(23.8, 36.2) | 12<br>.05 | 22.10<br>(16.1, 28.2) | 23.70<br>(17.7, 29.7) | 14<br>.97 | <.0001 |
|  | 15RM Chest<br>press (Kg) | 12.70<br>(6.5, 18.9) | 33.10<br>(26.9, 39.3) | 12<br><.0001 | 22.3<br>(16.52, 28.0) | 27.70<br>(21.74, 33.6) | 12<br>.09 | 22.80<br>(17.02, 28.5) | 24.00<br>(18.24, 29.8) | 14<br>.98 | <.0001 |
|  | 15RM knee<br>extension (Kg) | 12.90<br>(7.7, 18.1) | 27.30<br>(22.1, 32.5) | 12<br><.0001 | 15.6<br>(10.8, 20.4) | 23.20<br>(18.3, 28.1) | 12<br><.0001 | 15.10<br>(10.0, 20.2) | 16.20<br>(11.1, 21.2) | 14<br>.94 | <.0001 |

### Participants’ Demographics

The participants’ characteristics are presented in Table 1.

One Z-group participant dropped out. COVID-19 lockdown restrictions prevented complete post-testing for 3 G-group and 2 Z-group participants (Figure 1).

### Body anthropometrics

The interactions with BMI and WC, indices of body anthropometrics, were found to be statistically significant (Table 2). Post-hoc analysis revealed that while BMI did not change significantly in the training groups (P=.65 and P=1 for the G-group and Z-group, respectively), WC significantly reduced following training in the G and Z groups. In the G-group, there was a decrease of 2.042 cm (SE=.603, P=.0202), and in the Z-group, the decrease was 2.637 cm (SE=.603,.0014).

The difference in the reduction levels between the Z-group and G-group, 0.6 cm (SE=.852), was not statistically significant (P=.490). Unlike the intervention groups, in the C-group, BMI and WC both increased by .7143 kg/cm^2^ (SE=.217, P=.0256) and 1.657 cm (SE=.559, P=.056, marginal significance) between T1 and baseline testing (detailed at Table 2 and Figure 4).

**Figure 4.**
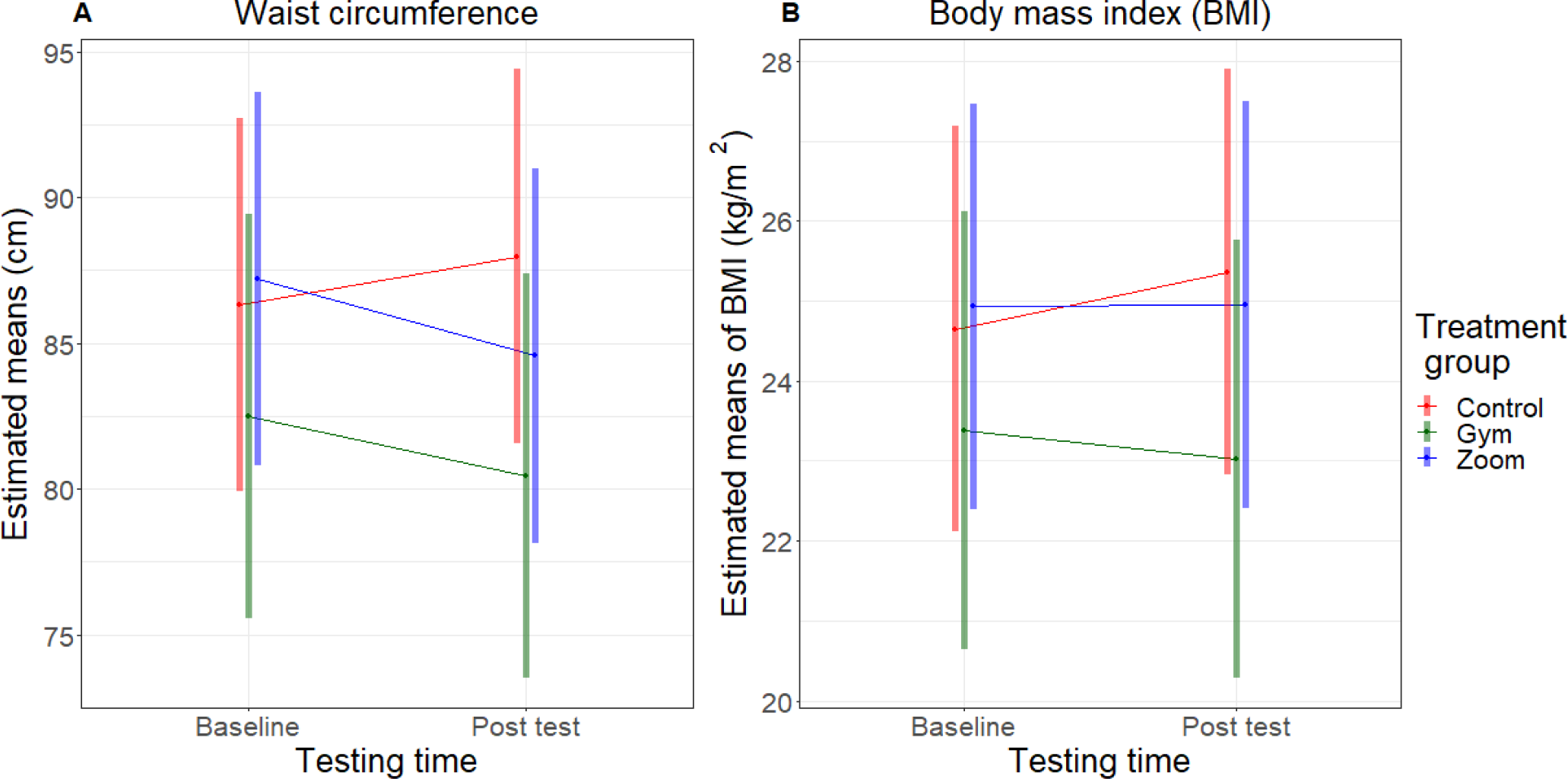
Body anthropometrics. The mean and 95-percent confidence interval of waist circumference (A) and body mass index (B) at baseline (T0) and post-testing (T1) for participants in the Gym Group (green), Zoom Group (blue), and Control Group (red).

### Blood Pressure

The interactions of the three Blood Pressure indices, SBP, and DSP, were also found to be statistically significant, and the P-values were <.0001, and .0001, respectively (Table 2 and Figure 5).

**Figure 5.**
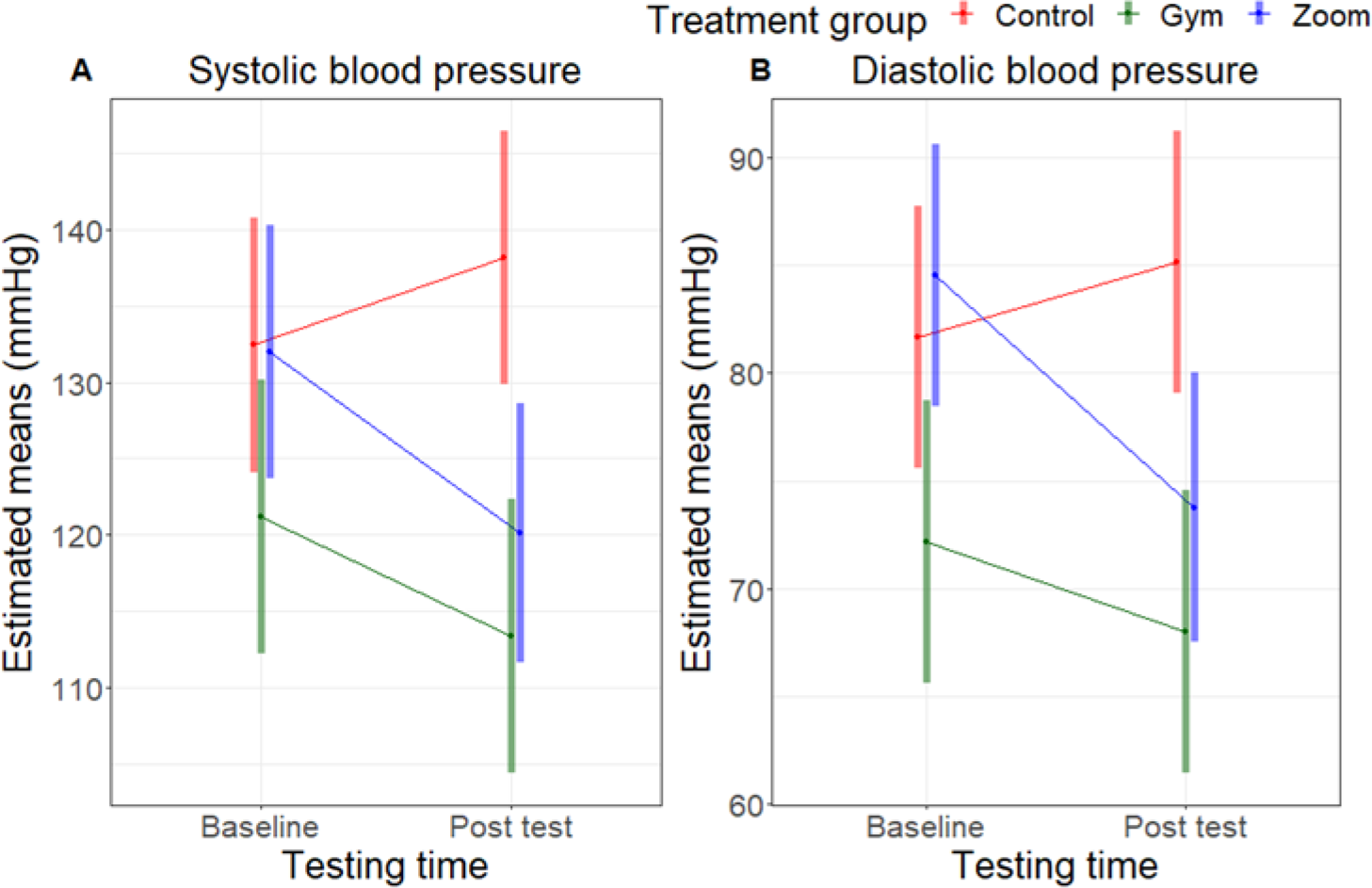
Blood pressure. The mean and 95-percent confidence interval of the systolic (A) and diastolic (B) blood pressure at baseline (T0) and post-testing (T1) for participants in the Gym Group (green), Zoom Group (blue), and Control Group (red).

The SBP decreased by 7.83 mmHg (SE=2.28, P=.018) and 11.91 mmHg (SE=2.27, P=.0001) in the G-group and Z-group, respectively, and the difference in the reduction level between G-group and Z-group, 4.074 mmHg (SE=3.223), was not statistically significant (P=.214). As for DBP, the decrease was 5.377 mmHg (SE=1.94, P=.086—marginal significance) and 11.104 (SE=1.93, P<.0001) in the G-group and Z-group, respectively, and the difference in the reduction level between the G-group and Z-group, 5.528 mmHg (SE=2.735), was statistically significant (P=.043).

In the C-group, the SBP increased by 5.714 mmHg (SE=2.11, P=.055—marginal significance) between baseline testing and T1 (Table 2, Figure 5).

### Aerobic Fitness

The LaLa test peak speed interaction was found to be statistically significant (P<.0001). Between T1 and T0, The LaLa test peak speed significantly increased (P<.0001) only in the Z-group, and the estimated mean peak speed increased by 2.243 km/hr (SE=.35). No significant changes were observed in the G-group and C-group (P=.99 and 0.92, respectively) (Table 2, Figure 6a).

**Figure 6.**
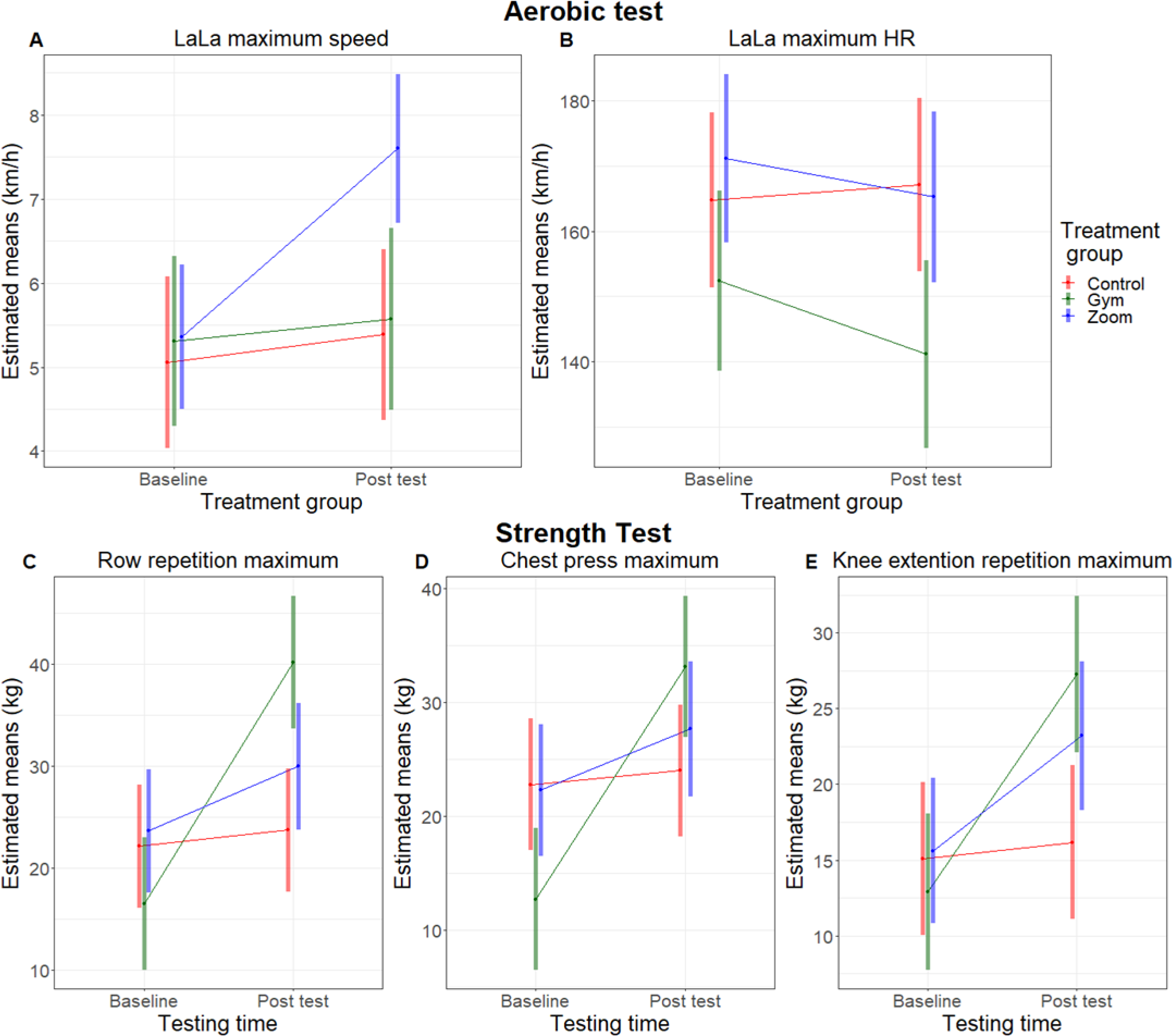
Aerobic and strength parameters: The mean and 95-percent confidence interval of the aerobic (A, B) and strength tests (C, D, and E) at baseline (T0) and post-testing (T1) for participants in the Gym Group (green), Zoom Group (blue), and Control Group (red).

In the case of the LaLa test Peak-HR, the interaction was marginally statistically significant (P=.0714). The drop in peak HR in the LaLa test was statistically significant in both intervention groups, lower by 11.27 bpn (SE=4.33, P=.0144) at T1 compared to T0 in the G-group and by 5.92 bpn (SE=3.91, P=.1413) in the Z-group. A slightly higher Peak HR of 2.27 bpn (SE=3.77) was measured in the C-group, but this increase was not statistically significant (P=.5516).

### Muscle Strength

In this group of variables, the interaction factor was found to be statistically significant for all variables, 15-RM-Row, 15-RM-Chest, and 15-RM-Knee, and all p-values were below .0001. The increases in 15-RM load in the strength tests were statistically significant at T1 in all three tests in G-group— an increase of 23.666 kg (SE=2.12P<.0001) for Row, 20.42 kg (SE=1.97, P<.0001) for Chest, and 14.375 kg (SE=1.25, P<.0001) for Knee extension. In the Z-group, we found a statistically significant increase only in two cases, 6.321 kg (SE=2.10, P=.50) for Row and 7.578 kg (SE=1.24, P<.0001) for Knee; in the third case, Chest, we found a marginally statistically significant increase at T1, 5.37 kg (SE=7.96, P=.091). In all the tests, no significant gains in the C-group were found [Table 2, Figure 6 (graphs B, C, and D)].

G-group’s gains were significantly greater than Z-group’s in the 15-RM-row by 17.346 kg (SE=2.986, P<.0001), and 15-RM knee-extension by 3.797 (SE=1.76, P=.005).

### Perceived Fitness Level

Perceived fitness was rated significantly higher in both G-group [Median=5 (Range=8) at T0 and Median=7 (Range=71.93) at T1] and Z-group [Median=5 (Range=6.5) at T0 and Median=6.5 (Range=5.5) at T1] (P=.00233, P=.0179, respectively). G-group showed a significantly larger increase in rating compared to Z-group (P=.0033). The control-group’s fitness ratings remained unchanged [T0: Median=5.5 (Range=5), T1: Median=5 (Range=7) p=.5807].

## DISCUSSION

This study examined the impact of two training methods—a real-time circuit tele-training program delivered on Zoom and a combined aerobic-resistance-training program at a community gym—comparing the outcomes of the two intervention groups with a those of a wait-list control group. The findings demonstrate that both training methods resulted in significant positive health-related gains in BP, body anthropometrics, strength, and aerobic fitness. Conversely, the control group exhibited no gains and even experienced deterioration in body anthropometrics and BP. Notably, the Zoom-training group (Z-group) showed more substantial improvements in DBP and aerobic capacity, while the gym-training group (G-group) exhibited greater gains in muscle strength.

Analyzing body anthropometrics, we observed no significant changes in BMI, but a significant similar reduction in WC in both exercise groups. WC, an indicator of visceral adiposity,^10,34^ is a substantial independent risk factor for cardiometabolic disease, making it a potentially more sensitive indicator than BMI.^10,35–37^ Our findings are consistent with previous records of reduced visceral fat volume in different populations with or without a reduction in BMI.^10,35^ Notably, our results emphasized a disadvantage of using BMI as an indicator of adiposity in CP populations.^5,36^

Compared to typically developing (TD) individuals, adults with CP have greater visceral, subcutaneous, and intermuscular adipose tissue; significantly smaller muscles; and lower bone-mineral density. Thus, individuals with CP could have greater visceral adiposity than TD individuals with the same BMI. Therefore, central adiposity estimated by WC^34^ is a more sensitive indicator than BMI for predicting cardiometabolic health risk in adults with CP.^36,38^

In contrast, the C-group displayed a significant rise in both BMI and WC between baseline (T0) testing and post-test (T1). This aligns with Van den Berg-Emon’s findings^18^ in a sports program for students with CP that resulted in a significant increase in fat mass in the control group but no changes in the exercising groups, emphasizing the importance of exercise in managing body composition.

Our study suggests that exercise may reduce visceral fat mass and increase muscle mass, impacting body composition and metabolic control in individuals with CP without reducing overall mass. Individuals with CP face an increased risk of being overweight, greater visceral adiposity,^39^ obesity, and obesity-related conditions.^36,37^ At baseline, 44% of our participants had at least one parameter indicating that they were overweight. Considering the association between reduced visceral adiposity and a decrease in cardiometabolic risk,^10,11^ our results indicate that exercise is effective in supplementary-weight management.

As for BP, both training groups demonstrated a significant and similar reduction in SBP, while the drop in DBP was significant in the Z-group and marginally significant in the G-group. Nevertheless, the mean reduction in SBP and DBP in both training groups were all above the clinically significant change for reducing morbidity risk.^11^ In contrast, the C-group displayed a marginally significant increase in SBP and a non-significant increase in DBP, all with clinical significance.^11^ Although in adults with CP the risk of hypertension is likely to increase with age,^6^ our study presents a decline in the C-group within months not within years as expected.^6^ In line with previous findings.^6,40^ At baseline, 56% of our entire sample had prehypertension, and 22% had hypertension. This is noteworthy, considering that among TD adults aged 21-34 years, only 3.3% of men and 2.8% of women had hypertension (63% of our participants were aged under 34 years). The prevalence in our findings aligns more with age groups between 45-54 and the 55-64 years. The improvements in BP following training are clinically significant, suggesting that both training methods can reduce the risk of disease and improve overall health. Notably, the BP of participants who did not train increased during this period. A 3 mmHg higher systolic and a 2.3 mmHg higher DBP translate to an estimated 12% increase in risk for chronic heart disease and a 24% increase in risk for stroke.^41^ Our findings bear important implications for health promotion, with mean reductions in SBP of 8 mmHg in the G-group and 12 mmHg in the Z-group, and 4.2 and 10.7 for DBP, respectively. Moreover, a meta-analysis involving one million people showed a linear relationship between decreased cardiovascular mortality risk and decreased BP to an SBP below 115 mmHg and a DBP below 75 mmHg.^42^ This reduction was demonstrated in 88% of our sample. While the greater DBP reduction in the Z-group may be due to the superiority of this training for reducing DBP, it may also be influenced by their baseline differences. The G-group sample had BP values slightly more within the normal range, whereas, the Z-group consisted of more prehypertensive participants, and greater benefits are documented for adults with prehypertension than for those with normal BP.^11^ The C-group’s deterioration in a short period of 3 months can be viewed in line with the general trend of weight gain and health deterioration during the Covid-19 epidemic.^43^ Our findings underscore the effectiveness of exercise in controlling body composition and BP, advocating for the using of WC and BP in health surveillance and assessment of health responses to training in adults with CP.

Turning to general fitness, both exercise groups reported significant improvements, with the G-group showing a more pronounced increase.

Aerobic capacity, measured by the LaLa test, significantly improved only in the Z-group. This improvement is attributed to the circuit training, which kept heart rate elevated throughout the sessions.^44^ Although a systematic review demonstrated strength and aerobic gains in the elderly following circuit training,^45^ to our knowledge, our study is the first to report aerobic capacity outcomes for such training in individuals with CP. Surprisingly, although aerobic training has been reported to improve aerobic capacity in individuals with CP, ^14^ the G-group did not exhibit a higher aerobic achievement. Despite incorporating recommended aerobic training intensity and frequency^12^ the lower duration of aerobic exercise in the G-group may be the reason. Nevertheless, significantly lower peak-HR measures in the G-group indicate an aerobic improvement. At baseline, the G-group’s aerobic peak-HR was 152.42 bpm, not reaching a HR of 180-bpm, which is considered a maximum effort for individuals with CP^46^; in contrast, HR reached 174.77 bpm in the Z-group and 168.83 bpm in the C-group. At post testing, the G-group reached an even lower HR, a drop of 11.27 bpm, while reaching about the same maximum speed. A similar incremental speed with a lower HR indicates an aerobic improvement. It should be mentioned that the Z-group completed the training at a significantly higher speed with a (non-significant) lower HR of 8-bpm; therefore, their aerobic gain may have been even greater. In contrast, the C-group displayed almost unchanged speed and peak HR. We suggest that the reason G-group did not exert maximum aerobic capacity is a lower ability to generate force rapidly in individuals with CP ^47^ rather than a lack of effort. Thus, maximum speed may be limited by reduced speed generation rather than aerobic capacity, at least for a portion of the participants. It has been suggested that power-training (involving rapid muscle contraction) is effective in improving the ability of individuals with CP to produce higher walking speed.^48^ These results suggest potential advantages of circuit training in individuals with CP, improving limited speed production since movement forces are rapidly generated in training.

Strength tests indicate significant improvements in MGF in both exercise groups, with the G-group demonstrating superior gains and with no significant improvement in the C-group. The positive outcomes align with existing literature on the effectiveness of resistance training in individuals with CP.^15–17^

### Limitations

The implications of this research should be considered given its limitations. One limitation is the small sample size. The study might be underpowered for the Z-group, as two participants missed part of their measures, and is most certainly underpowered for the G-group, as only 9 of 12 participants completed all tests because the pandemic interfered with the post-testing. Another limitation is that we didn’t utilize a randomized design for this study. In addition, although the training was planned in detail, there may be flaws to its design.

Specifically, aerobic training duration in the G-group was shorter than recommended and frequency was the minimum required for improvement but less than optimal.^11–12^ Finally, the resemblances between the strength test and exercise in the G-group, using testing machines, should be acknowledged.

### Conclusions

Despite the limitations, this study contributes valuable insights into the positive effects of exercise for ambulatory adults with spastic CP. Both training programs demonstrated significant reductions in health-risk factors, emphasizing the potential for exercise to enhance health in this population. The comparative analysis suggests that circuit training may be more effective in improving aerobic and BP measures, while resistance training is better for enhancing muscle strength. Additionally, the study supports utilizing such noninvasive measures as WC and BP for health surveillance and measuring exercise health benefits. This study presents two effective training programs that could increase participation in physical training and assist in managing health risks in adults with CP.

## Data Availability

All data produced in the present study are available upon reasonable request to the authors

## ACKNOWLEDGMENTS

The authors thank all participants for taking part in this study. They also thank Yuliya Timoshevsky, for solving unexpected problems and for assisting with data collection and logistics. The authors would like to thank the physical therapists Yochi Cohen, Efrat Rozenberg, and Tzofit Zmora, and coaches Hades, Shlomy, Dema, and Yanir for their parts in testing or coaching in this study. The authors are grateful for the cooperation with the owner and staff at Lezoz gym in Jerusalem for allowing us to use the facilities free of charge. We also thank the team of the Sports Center of the Direction in Be’er Sheva and Mayor Rubik Danilovich for support and use of the facilities.

## FUNDING

This study was supported by the Israeli National Insurance Research Fund.

## ROLE OF THE FUNDING SOURCE

The funders played no role in the design, conduct, or reporting of this study.

## ETHICS APPROVAL

The study protocol was approved by the Medical Ethics Committee of Shamir Medical Center (Assaf Harofeh), affiliated with Tel-Aviv University, Israel.

## CLINICAL TRIAL REGISTRATION

This trial registration was prospectively registered in the in the Israeli database MOH_2018-02-14_002188.

## DATA AVAILIBILITY

The data will be provided by the researchers upon request.

## DISCLOSURE

The authors completed the ICMJE Form for Disclosure of Potential Conflicts of Interest and reported no conflicts of interest.

## SUPPLEMENTARY MATERIAL

### The gym-group (G-group) and Zoom-group (Z-group) interventions

Participants in both groups were instructed not to alter their routine physical activity (PA) during this study.

The G-group training took place in Kivunim, a community center in the city of Beer-Sheva, Israel. The Z-group trainers and participants engaged in real-time Zoom circuit tele-training in their homes.

Coaches participated in a preliminary seminar. Midway through the program, participants were sent an online feedback questionnaire (adjusted with permission from Schwartz and colleagues^1^).

Conducted with groups of 6-8 participants, training sessions were held twice a week on non-consecutive days for a total of 32 sessions and were one hour long. Each session began with a 5-10 minute warm-up and concluded with a 10-minute cool-down and stretching.

Both training programs were individually tailored, offering several alternatives with varying degrees of difficulty and adjustments. Special attention was given to ensuring an independent and safe transition between machines (g-Group) and between exercise positions (in both groups), with participants gradually achieving independence in performance and adjustment. Participants were encouraged to maintain an adequate exercise form within their personal limits.

### Adjustment Period

The first 2-4 weeks were designated as an adjustment period in which the protocol was established personally, and participants became familiar with it. Additionally, participants practiced self-identifying their own aerobic effort using the talk-test^2^ and identifying resistance-exercise load effort using the 2-for-2 principle.^3^

The talk-test is a practical and reliable tool for prescribing and monitoring exercise intensity and is applicable to various modes of exercise. The test is based on the rationale that a higher exercise intensity requires deeper breathing, making talking harder.^2^

According to the 2-for-2 rule, an overload is recommended if, in the last set of a resistance exercise, participants complete two or more repetitions beyond their assigned repetition goal for any exercise for two consecutive workouts.^3^

### Progression

Participants were prompted to report when they recognized that they could update an exercise using the talk-test for aerobic effort and the 2-for-2 principle for recognizing resistance effort. The protocols were then collaboratively assessed, adjusted, and progressed by the participants and staff approximately every 3 weeks. Participants were also asked to report the initiation or aggravation of musculoskeletal pain during exercise, at which point the training was adjusted to avoid pain.

### G-group: aerobic and resistance training program at a community gym

The G-group training program was guided by two coaches, a physical therapist, and an assistant, who provided guidance, assistance, feedback, and encouragement.

#### Training Structure

Participants trained in two random groups. One group began with strength training for about 30-40 minutes, while the second group started with aerobic training for about 20 minutes. The groups then swapped places and continued with the second part.

#### Aerobic Training

##### Type

All programs included treadmill walk/run exercise, some of which were included throughout the training, while others, according to preference, were combined with an additional type of exercise including a stationary cycling bike, cross-training machine, stepper, or an Adaptive Motion Trainer. *<u>Intensity:</u>* In the talk-test, the goal was to select an intensity where speech became somewhat uncomfortable, up to an intensity where participants could “just respond to the conversation”.^4^ Additionally, using HR monitoring, the aerobic HR target training zone began with a 40-60% Heart Rate Reserve (HRR) in the adjusting period, building up to a 60-80% HRR training zone.

The target HR was calculated using the Karvonen formula,^5^ assuming HR-max of 180 bpm^6^ and measuring resting HR (RHR). The formula is calculated as follows: Target HR=((HR-max−RHR)×40%−80%)+RHRTarget HR=((HR-max−RHR)×40%−80%)+RHR Initially, participants were instructed to walk at the mean speed calculated from their 6MWT (T0) and then build up to the target range (Tables 1a and 1b), to the extent tolerable to them. Participants could reduce intensity to the lower HR target zone or to a comfortable zone for a short duration if necessary.

**Table 1a:** Aerobic Training - weeks 1-8.

| Table 1a: Aerobic Training – weeks 1-8 |  |  |  |  |
| --- | --- | --- | --- | --- |
|  | Acclimation/familiarization/establishing the personal program and machines/ familiarization with Talk test and target HR |  | Building up endurance |  |
| Volume of training / Week | Week 1-2 | Week 3-4 | Week 5-6 | Week 7-8 |
| Target HR | HRR 40% | HRR 40-60% | 50-65% HRR | HRR 55-70% |
| Duration | Familiarization: Aiming to complete 10 min with active intervals on TM. Experiencing additional exercise on machines. | 10-20 min with intervals: high / lower for adaptation | 10-20 min with intervals: high/ lower for adaptation | 15-20 min |
| Intervals | 2/2: 2 minutes comfortable walking Speed (CWS) / 2 minutes target HR | 2/2 Combined with continuous higher tolerated target zone intensity | Aiming for the higher target zone, combining 2-minute intervals of the lower target zone according to tolerance. |  |
| Talk test | Speech is comfortable, but may become somewhat difficult, requiring short intervals. | Aiming for the higher zone of comfortable talking | Aiming for the higher zone of comfortable talking for longer periods | Speech is comfortable, but may become somewhat difficult |

**Table 1b:** Aerobic Training - weeks 9-16.

| Table 1b: Aerobic Training – weeks 9-16 |  |  |  |  |
| --- | --- | --- | --- | --- |
| Building up endurance |  |  |  |  |
| Volume of training / Week | Week 9-10 | Week 11-12 | Week 13-14 | Week 15-16 |
| Target HR | HRR 55-70% | HRR 60-80% | HRR 60-80% | HRR 65-80% |
| Duration | 20 min | 20 min | 20 min with intervals: high / lower for adaptation | 20 min |
| Intervals | Aiming for the higher target zone, combining 2-minute intervals of lower target zone according to tolerance |  | Aiming to reduce the lower speed intervals | Aiming to reduce the lower speed intervals |
| Talk test | Speech is comfortable, but may become somewhat difficult | Speech is somewhat difficult- participant can only respond to conversation |  |  |

#### Resistance Training

The resistance training included 7-12 exercises of major muscle groups. For the exercise load and progression protocol, see Table 2a and 2b.

**Table 2a:** Progressive Resistance Training – weeks 1-8.

| Table 2a: Progressive Resistance Training – weeks 1-8 |  |  |  |  |
| --- | --- | --- | --- | --- |
|  | Acclimation/familiarization/establishing the personal program and machines/ familiarization with the feeling of high-effort training and the two-for-two principal. |  | Muscle endurance |  |
| Volume of training / Week | Week 1-2 | Week 3-4 | Week 5-6 | Week 7-8 |
| Number of sets | 1 | 2 | 2 | 2 |
| Intervals between sets | 30-90 sec | 30-90 sec | 30-90 sec | 30-90 sec |
| Resistance / Number of repetitions | Minimum (beginners) | Gradual build towards 12-15 RM | 15 RM | 12-15RM |

**Table 2b:** Progressive Resistance Training – weeks 9-16.

| Table 2b: Progressive Resistance Training – weeks 9-16 |  |  |  |  |
| --- | --- | --- | --- | --- |
|  | Muscle endurance and hypertrophy |  | Muscle endurance |  |
| Volume of training/ Week | Week 9-10 | Week 11-12 | Week 13-14 | Week 15-16 |
| Number of sets | 3 | 2 | 2-3 | 2-3 |
| Intervals between sets | 30-90 sec | 30-90 sec | 30-90 sec | 30-90 sec |
| Resistance / Number of repetitions | 12-15 RM | 10-12 RM | 10-12 RM | 10-12 RM |

### Z-group: real-time circuit tele-training delivered on Zoom

Before the exercise sessions, participants received links to two 10-minute videos. One video introduced the staff and included information regarding wanted and unwanted sensations during and after training and safety guidelines. The other video contained a short version of the session exercise at three different exercise difficulty levels.

The first session was a one-on-one Zoom session in which the exercise and computer location were set for full-body observation. In addition, the initial tailored exercises program was chosen and emailed to the participants. Halfway through the program, the participants were offered another one-on-one meeting. The group-training was guided once a week by a coach and once by a physical therapist. The Zoom sessions were recorded and monitored, and feedback was sent to participants. The training was structured as circuit resistance training. The program comprised ∼10 exercises of 6–15 repetitions, targeting different muscle groups. Each exercise was performed for a duration of 20 seconds and repeated twice before moving on to the next exercise. The trainees moved quickly from one exercise to the next with a 10-second recovery with the same exercise starting position and a 20-second recovery when a different starting position was required.

#### Load and Progression

Participants were instructed to perform a maximum number of repetitions in 20 seconds. The target was to complete two sets of 6-15 repetitions. When participants were unable to complete at least 6 repetitions of an exercise, when they exceeded 15 repetitions, or if they felt at the end of the second set that they could go on with this exercise for two or more repetitions (the 2-for-2 principle), the exercise was adjusted.

## Notes

### Competing Interest Statement

The authors have declared no competing interest.

### Clinical Trial

The trial is registered in the Israeli database MOH_2018-02-14_002188

### Author Declarations

Ethics committee of Shamir Medical Center (Assaf Harofeh), affiliated with Tel Aviv University, Israel (approval number 0223-17-ASF) gave ethical approval for this work.

